# Quantitative PET imaging and modeling of molecular blood-brain barrier permeability

**DOI:** 10.1101/2024.07.26.24311027

**Authors:** Kevin J. Chung, Yasser G. Abdelhafez, Benjamin A. Spencer, Terry Jones, Quyen Tran, Lorenzo Nardo, Moon S. Chen, Souvik Sarkar, Valentina Medici, Victoria Lyo, Ramsey D. Badawi, Simon R. Cherry, Guobao Wang

## Abstract

Blood-brain barrier (BBB) disruption is involved in the pathogenesis and progression of many neurological and systemic diseases. Non-invasive assessment of BBB permeability in humans has mainly been performed with dynamic contrast-enhanced magnetic resonance imaging, evaluating the BBB as a structural barrier. Here, we developed a novel non-invasive positron emission tomography (PET) method in humans to measure the BBB permeability of molecular radiotracers that cross the BBB through different transport mechanisms. Our method uses high-temporal resolution dynamic imaging and kinetic modeling to jointly estimate cerebral blood flow and tracer-specific BBB transport rate from a single dynamic PET scan and measure the molecular permeability-surface area (PS) product of the radiotracer. We show our method can resolve BBB PS across three PET radiotracers with greatly differing permeabilities, measure reductions in BBB PS of ^18^F-fluorodeoxyglucose (FDG) in healthy aging, and demonstrate a possible brain-body association between decreased FDG BBB PS in patients with metabolic dysfunction-associated steatotic liver inflammation. Our method opens new directions to efficiently study the molecular permeability of the human BBB *in vivo* using the large catalogue of available molecular PET tracers.

---

The blood-brain barrier (BBB) regulates molecular exchange between the blood and the brain. The BBB not only comprises a structural barrier that tightly restricts blood-to-brain solute diffusion, but also numerous molecular transport systems that support nutritive transport for brain function (Figure 1).^1–3^ BBB dysfunction is accordingly often associated with a change in BBB permeability, for example, through loss of blood solute filtration during BBB breakdown or through altered BBB transport systems.^1–3^

**Figure 1.**
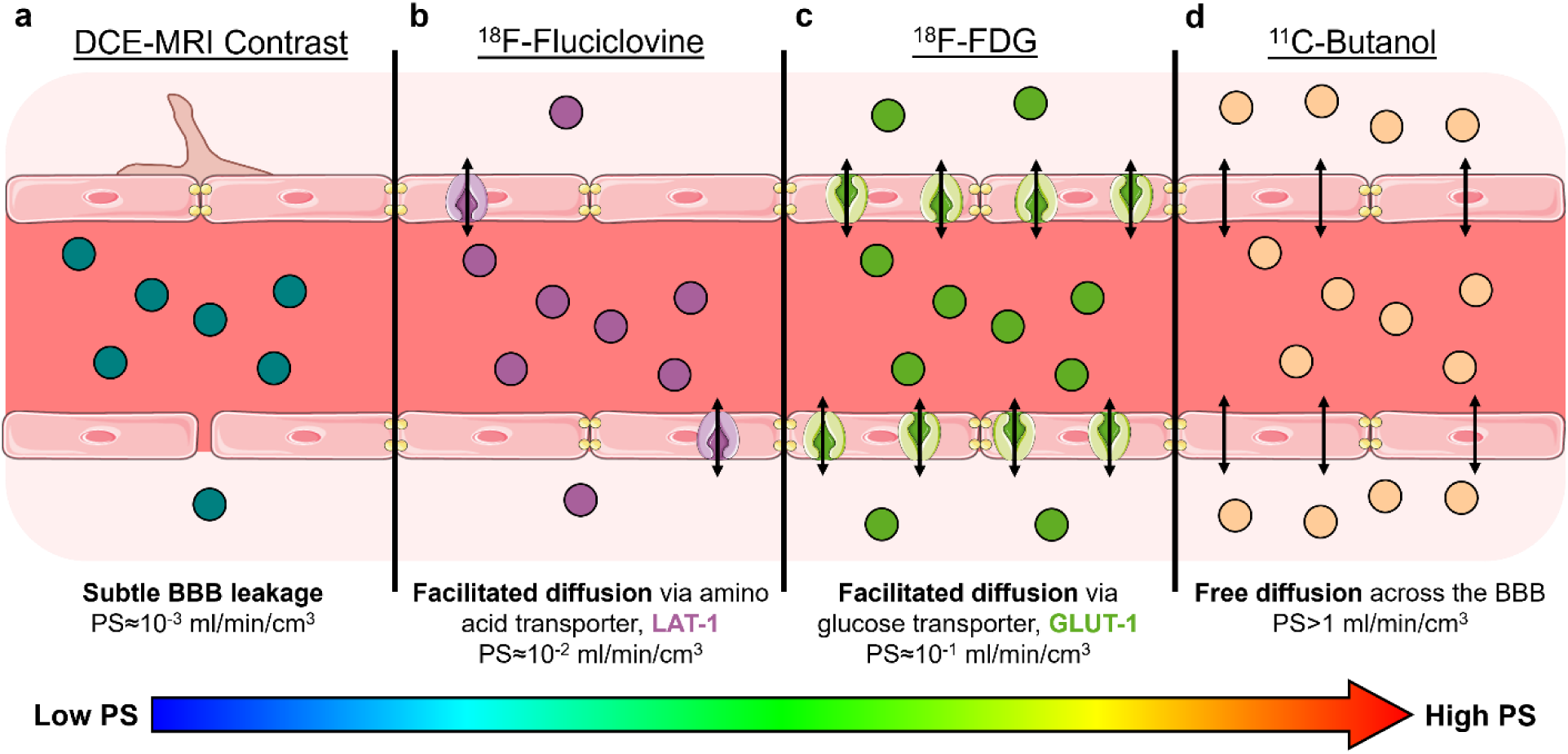
The blood-brain barrier (BBB) is commonly treated as a structural barrier (e.g., tightly lined endothelial cells, tight junction proteins, astrocyte end feet) but also comprises various transport systems and mechanisms associated with molecular BBB permeability^1–3^ that can be imaged using the proposed method with many available PET radiotracers. **a,** Current methods of BBB permeability imaging mainly evaluate its structural integrity with gadolinium contrast-enhanced dynamic magnetic resonance imaging (DCE-MRI), where an increase in DCE-MRI measures of permeability reflect non-specific BBB leakage. The BBB permeability-surface area (PS) product with gadolinium DCE-MRI is on the order of 10^-3^ ml/min/cm^3^.^8^ **b**, ^18^F-fluciclovine is a radiolabeled analogue of an essential amino acid thought to cross the BBB via facilitated diffusion through the large neutral amino acid transporter 1 (LAT1; purple transporter),^53^ which we show has a BBB PS on the order of 10^-2^ ml/min/cm^3^. **c**, ^18^F-fluorodeoxyglucose (FDG) is the ubiquitous glucose analogue that mainly crosses the BBB via facilitated diffusion through glucose transporter 1 (GLUT1; green transporter)^54^ and a BBB PS on the order of 10^-1^ ml/min/cm^3^. **d**, ^11^C-butanol is a lipophilic radiolabeled alcohol that freely diffuses across the BBB^16,23^ with a BBB PS on the order of >1 ml/min/cm^3^.

Neuroimaging of BBB permeability has been instrumental in assessing BBB dysfunction as a hallmark of many neurological and systemic disorders.^1,4^ However, current *in vivo* methods mainly focus on assessing the BBB as a structural barrier. Dynamic contrast-enhanced (DCE) magnetic resonance imaging (MRI) uses gadolinium contrast agents to assess the structural integrity of the BBB under the assumption that these agents do not effectively cross a normal BBB and accordingly have very low normal BBB permeability.^5^ The permeability-surface area (PS) product is a specific kinetic measure of BBB permeability^6,7^ with an order of magnitude of 10^-3^ ml/min/cm^3^ for gadolinium MRI contrast agents.^8^ DCE-MRI measures of BBB permeability have been shown to increase with aging,^8^ cognitive impairment,^9^ and Alzheimer’s disease due to subtle BBB leakage.^10^

BBB transport also occurs through molecular transporter mechanisms but measuring the associated permeability remains less explored in humans *in vivo*.^1–3^ We hypothesize that measuring the BBB PS of PET molecular radiotracers may open new opportunities to probe the human BBB at the molecular level. There are numerous neuroimaging PET radiotracers,^11^ each with distinct molecular BBB permeability properties stemming from their individual BBB transport mechanisms. For example, the ubiquitous glucose metabolism radiotracer ^18^F-fluorodeoxyglucose (FDG) crosses the BBB mainly via glucose transporter 1 (GLUT1) with a PS on the order of 10^-1^ ml/min/cm^3^.^12,13^ Thus, ^18^F-FDG PET has the potential to assess both cellular metabolism and molecular BBB function.

However, the BBB permeability of radiotracers has received limited attention in part due to the lack of efficient imaging tools. Currently, two PET scans with two radiotracers are required to measure the BBB permeability of a PET tracer,^14–16^ one for measuring BBB transport rate of the target tracer and the other for measuring cerebral blood flow (CBF) using a highly-extracted flow radiotracer (e.g., ^11^C-butanol^16,17^). This approach has faced limited use because dual-tracer protocols are costly, demand extensive infrastructure, and are challenging to execute in part due to the short half-lives of many flow radiotracers. Furthermore, conventional PET scanners have short axial coverage and limited spatial resolution to non-invasively obtain an accurate arterial input function for tracer kinetic analysis,^18^ necessitating invasive arterial blood sampling.^19^ These factors collectively contributed to the limited exploration of the BBB PS of radiotracers despite the potential ability to probe the molecular permeability of the human BBB *in vivo* using the large existing catalogue of molecular PET tracers.

Here, we developed a novel non-invasive multiparametric PET method to image and quantify the molecular BBB PS of radiotracers without a flow tracer PET scan. Our approach is enabled by the recent advent of high-sensitivity long axial-field-of-view PET that provides both high-temporal resolution (HTR) dynamic brain imaging and arterial blood pool imaging.^20,21^ Using advanced HTR kinetic modeling, the proposed method jointly estimates CBF and tracer-specific BBB transport rate K_1_ from a single HTR dynamic scan, which in turn provides quantification of the molecular PS of the radiotracer. We tested this method across three very different PET radiotracers and evaluated its application in healthy aging and in patients with metabolic dysfunction-associated steatohepatitis (MASH).

## Results

### High-temporal resolution dynamic PET enables joint imaging of CBF and tracer-specific BBB transport rate from a single dynamic scan

The ultra-high sensitivity of total-body PET scanners^20,21^ enables high temporal resolution dynamic brain PET imaging (e.g., 1 to 2 s per frame) compared to conventional PET scanners, which are practically limited to 5 to 20 s temporal resolution. Their extended axial field-of-view also allows a fully quantitative image-derived input function to be obtained from a major blood pool (e.g., the ascending aorta) while synchronously imaging the brain, obviating the need for invasive arterial blood sampling with minimal delay and dispersion effects^22^ for PET kinetic analysis (Figure 2a). The importance of HTR imaging to sample the radiotracer’s rapid first pass in the blood pool with high fidelity (Figure 2b) is illustrated in ascending aorta data shown at 1 s, 5 s, and 10 s frame durations.

**Figure 2.**
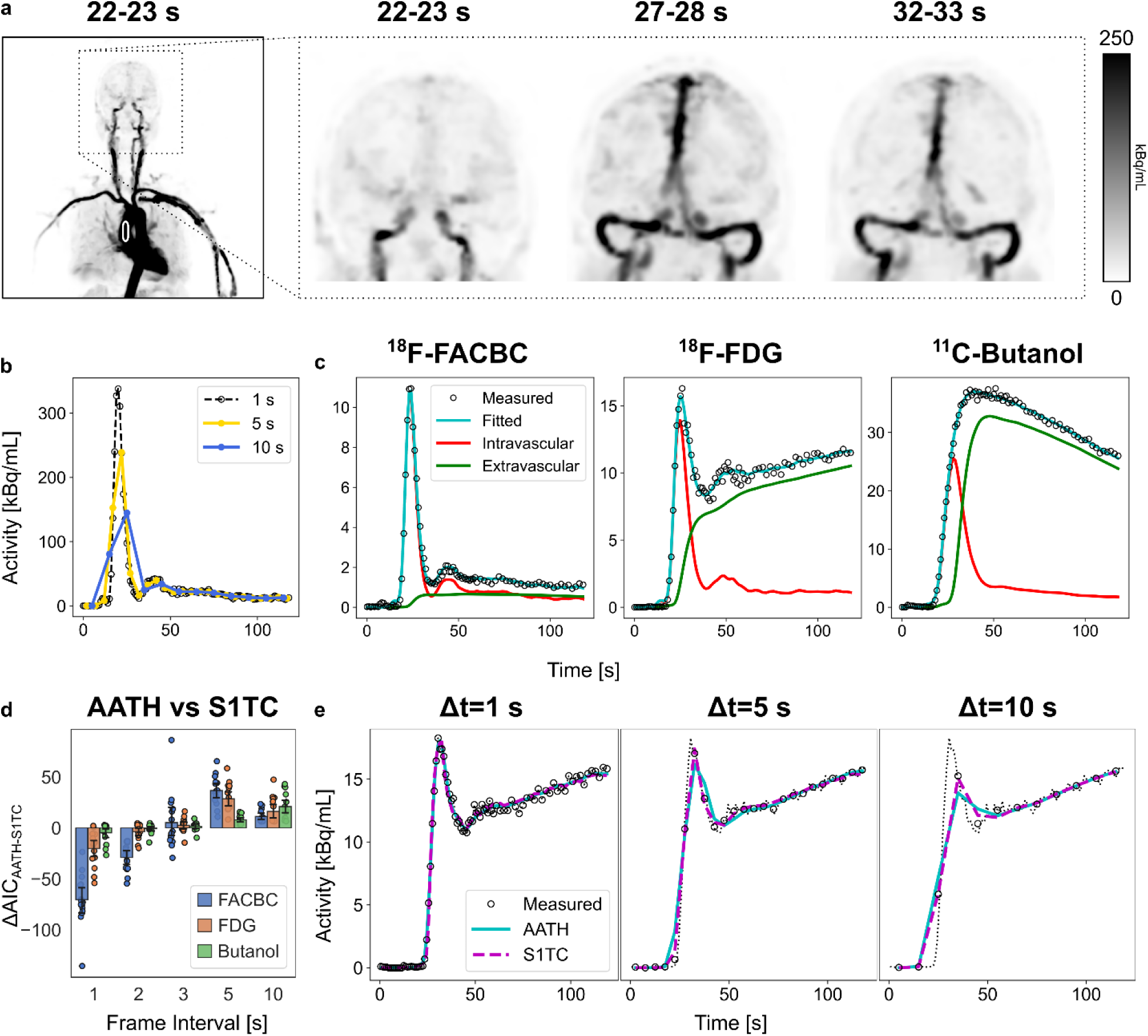
Total-body PET-enabled high-temporal resolution (HTR) dynamic imaging and kinetic modeling for non-invasive quantification of molecular blood-brain barrier (BBB) transport kinetics. **a**, Maximum-intensity coronal projections of three 1-s frame dynamic reconstructions (kBq/mL). The extended axial field of view allowed non-invasive measurement of the image-derived input function from the ascending aorta (white outline). **b**, A representative image-derived input function illustrating the importance of high temporal resolution to accurately sample the rapid transport of tracer through the blood pool. **c**, Representative fits to high temporal resolution time-activity curves of ^18^F-fluciclovine (FACBC), ^18^F-fluorodeoxyglucose (FDG), and ^11^C-butanol using the adiabatic approximation to the tissue-homogeneity (AATH) model. Fitted curves (teal) were decomposed into their intravascular (red) and extravascular tissue (green) distributions according to the AATH model. **d**, The difference in Akaike Information Criterion (AIC) between the AATH and the standard one-tissue compartment (S1TC) model time-activity curves frame averaged to different intervals. The AATH model was preferred over the S1TC for 1 to 2 s HTR frame intervals (negative AICs), but not for the 3, 5, 10 s intervals, illustrating the importance of total-body PET in enabling the non-invasive single-tracer BBB PS imaging method. **e**, Representative AATH and S1TC fits to an FDG time activity curve (dashed black line: original) in the grey matter at 1, 5, and 10 s frame intervals, with progressively poorer early peak fitting at greater frame intervals.

Standard compartmental models assume that a tracer instantaneously crosses and uniformly mixes in local blood vessels, neglecting the finite transit time required for the tracer to traverse the blood volume at a rate equal to blood flow. This assumption is reasonably valid for dynamic PET data at standard temporal resolutions (e.g., 5 to 20 s) but is less suitable for modeling HTR data as indicated by very early studies^23,24^ in the 1990’s and our initial studies.^25,26^ Hence we combined HTR dynamic imaging on total-body PET with advanced kinetic modeling approaches to resolve the tracer’s rapid vascular transit via CBF as well as its extravascular transport. Despite their application in dynamic contrast-enhanced MRI and CT,^27,28^ HTR kinetic modeling has historically received little attention in PET^23,24^ until recently^25,26,29^ due to the limited count levels of conventional PET scans. Here, we used the adiabatic approximation to the tissue homogeneity (AATH) model^30^ on the first two minutes of HTR dynamic PET scans (60×1 s frames then 30×2 s frames) to jointly estimate CBF and tracer-specific BBB transport rate K_1_ from a single dynamic HTR scan. We tested this method on total-body early dynamic PET scans of three different radiotracers (^18^F-fluciclovine, ^18^F-FDG, and ^11^C-butanol; Figure 1). The AATH model accurately fit the measured brain TACs for all investigated radiotracers at HTR (Figure 2c). By the Akaike Information Criterion (AIC),^31^ the AATH model better fit the measured data at 1 to 2 s HTR, but the standard one-tissue compartment model was favoured at 5 to 10 s temporal resolution (Figure 2d), highlighting the need for HTR to enable our method. Practical identifiability analysis showed that CBF and K_1_ were exceptionally identifiable, with <5% parameter bias and <15% error standard deviation (Supplementary Table 1; Supplementary Figure 1).

### Molecular BBB PS Differs Across PET Radiotracers

The joint estimation of CBF and BBB K_1_ enables calculation of the tracer extraction fraction (E; E = K_1_ / CBF) and the BBB PS using

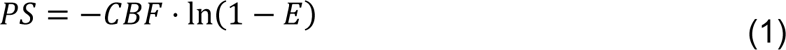

based on the Renkin-Crone equation.^32,33^ The practical identifiability of PS also remained excellent (absolute mean error < 5%, standard deviation < 12%; Supplementary Table 1) using real data and for simulated extraction fractions from 2.5 to 75% (Supplementary Figure 1). Note that existing dual-tracer methods with conventional PET scanners required an additional flow-specific radiotracer to separately estimate CBF to calculate E and PS. To demonstrate that our method can quantify molecular BBB PS of different PET tracers, we analyzed early dynamic PET scans of fifteen subjects scanned with either ^18^F-fluciclovine, ^18^F-FDG, or ^11^C-butanol on the uEXPLORER total-body PET/CT system.^21^ These radiotracers were selected to span a wide range of molecular BBB PS values according to their known range of BBB transport rate K_1_ values.^12,16,34^

We found that BBB PS greatly differed between the investigated PET tracers (Figure 3a), with BBB PS on the order of 10^-2^, 10^-1^, and >1 ml/min/cm^3^ for ^18^F-fluciclovine, ^18^F-FDG, and ^11^C-butanol, respectively. The mean ± standard deviation whole-brain PS of ^18^F-fluciclovine and ^18^F-FDG were 0.016±0.003 and 0.132±0.010 ml/min/cm^3^, respectively, while that of ^11^C-butanol was indeterminately high due to its free apparent diffusion across the BBB (i.e., Eq. (1) is indeterminate when E is 1).^16^ The radiotracers with higher apparent BBB PS had greater extravascular distribution according to the area underneath the intravascular and extravascular subcomponents of the fitted curve (Figure 2c) in part due to their differing BBB permeabilities. Regional BBB PS was significantly different between ^18^F-fluciclovine and ^18^F-FDG (P<0.001) in the grey matter, white matter, and cerebellum. Accordingly, we observed significant differences in regional K_1_ and E between ^18^F-fluciclovine, ^18^F-FDG, and ^11^C-butanol (P<0.001) but not for CBF (P>0.50), indicating that our method appears to estimate CBF consistently between PET tracers. Median (interquartile range, IQR) E across all brain regions were 4.7% (IQR: 3.7 to 5.1%), 32.6% (IQR: 29.9 to 38.4%), and 100% (IQR: 94.7 to 100%) for ^18^F-fluciclovine, ^18^F-FDG, and ^11^C-butanol, respectively.

**Figure 3.**
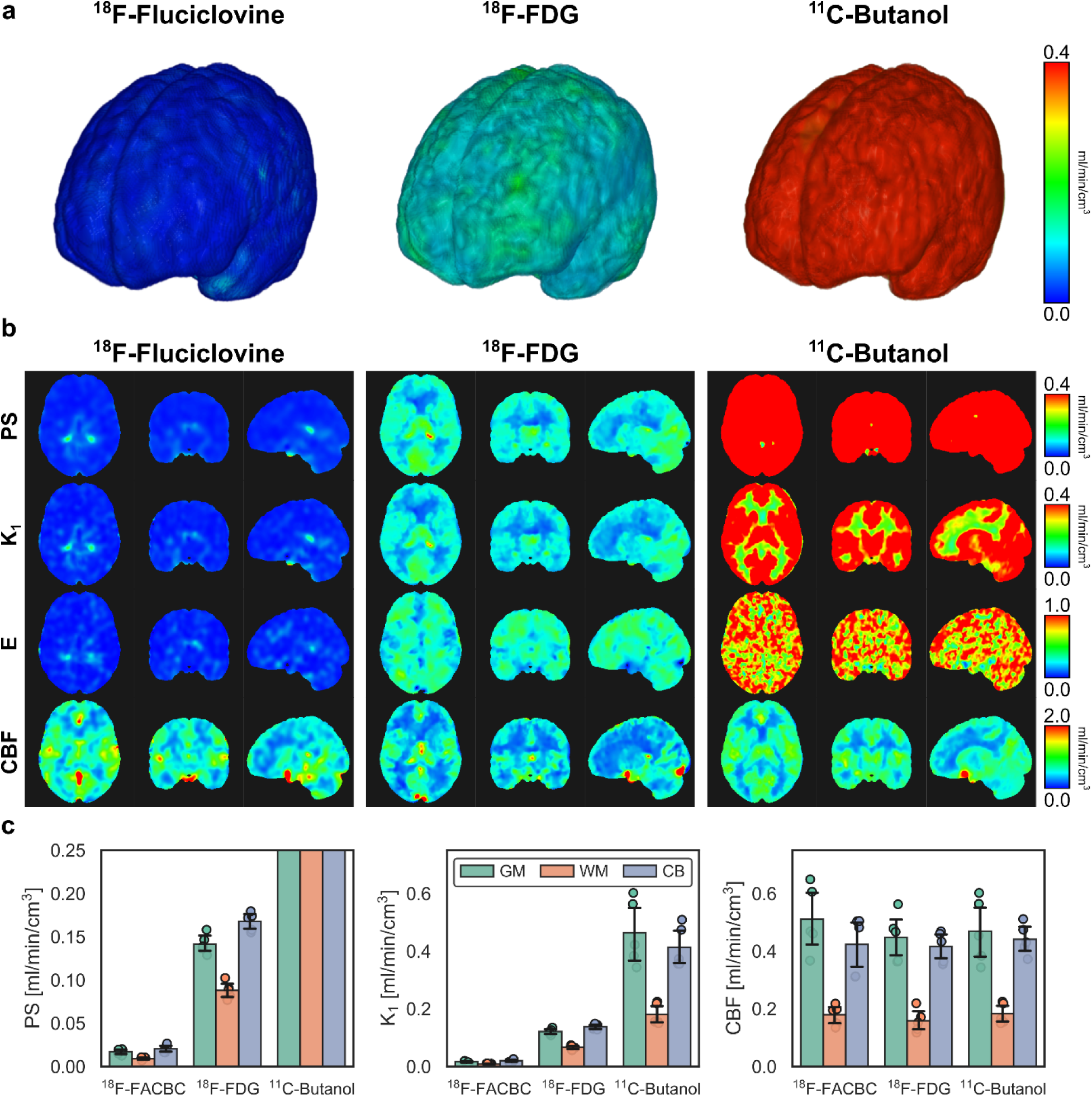
Imaging and quantifying the molecular blood-brain barrier (BBB) permeability-surface area (PS) product of three PET radiotracers. **a**, 3D renderings of molecular BBB PS maps with ^18^F-fluciclovine (FACBC), ^18^F-fluorodeoxyglucose (FDG), and ^11^C-butanol illustrating the spectrum of molecular BBB PS across PET radiotracers. **b**, Orthogonal slices of tracer PS, K_1_, extraction fraction (E), and cerebral blood flow (CBF) for each tracer. Parametric images were aligned to the MNI152 space for visualization. Of note, the hot spots in the ^18^F-fluciclovine PS and K_1_ maps are near the choroid plexus, reflecting the higher inherent permeability of the blood-cerebrospinal fluid (CSF) barrier compared to the BBB, while for the butanol PS map, the cold spots are at CSF pools in the ventricles. **c**, Regional quantification at the grey matter (GM), white matter (WM), and cerebellum (CB) shows substantial differences in PS and K_1_ between tracers (averaged across five subjects per tracer) while CBF appears comparable between tracers. PS, K_1_ and CBF are in units of ml/min/cm^3^.

Regional differences in BBB PS (Figure 3b) were observed in parametric imaging, particularly for ^18^F-FDG, which had substantial grey-white matter contrast and elevated PS in the cerebellum, both of which were corroborated by regional kinetic analysis (Figure 2c). For ^18^F-fluciclovine, BBB PS, K_1_, and E overall had small values with the exception of hot spots seen at the blood-cerebrospinal fluid (CSF) barrier of the choroid plexus, which is known to be inherently permeable in part due to fenestrations at its vasculature.^35^ In contrast, ^11^C-butanol had very high PS values and many voxels had E values of 100% (Figure 3b). The cold spots in the ^11^C-butanol PS map were at ventricular pools of CSF leading to an underestimation of regional PS. CBF maps appeared visually comparable between PET tracers, though the prominence of veins (e.g., sagittal sinus) appeared to decrease for radiotracers with higher PS possibly due to their higher tissue extraction leaving less radiotracer concentration cleared through the venous circulation.

### BBB PS of ^18^F-FDG Decreases in Healthy Aging

To demonstrate a potential application of the proposed method, we studied the association between age and the BBB permeability of ^18^F-FDG in healthy subjects. BBB breakdown and decreased glucose metabolism have been associated with aging,^8,36,37^ but it remains unclear whether BBB permeability changes at the molecular level in aging brains. We analyzed thirty-four healthy subjects in their mid-20s to late-70s (mean: 51±13 years) who underwent total-body dynamic FDG-PET. Regional HTR kinetic analysis of cortical grey matter showed that FDG BBB PS was significantly associated with age (P<0.001) (Figure 4). Linear regression predicted a cohort decrease in cortical FDG BBB PS of 8.58×10^-4^ ml/min/cm^3^ per year of older age, corresponding to a 0.23% decrease in cortical FDG BBB PS per year. A decreasing trend with age was similarly seen for FDG BBB PS in white matter and cerebellum but associations only approached significance (P=0.08 and P=0.05, respectively). Other demographic factors such as sex and body mass index (BMI) were not significantly associated with FDG BBB PS at other brain subregions in our investigation. BBB transport rate K_1_ showed significant associations with age in all studied brain regions (Figure 4; P<0.05), likely related to the joint decrease of FDG BBB PS and CBF with age (Figure 4). Linear regression predicted that K_1_ decreased at a rate of 0.27%, 0.15%, and 0.15% per year in the cortical grey matter, white matter, and cerebellum, respectively, in our cohort.

**Figure 4.**
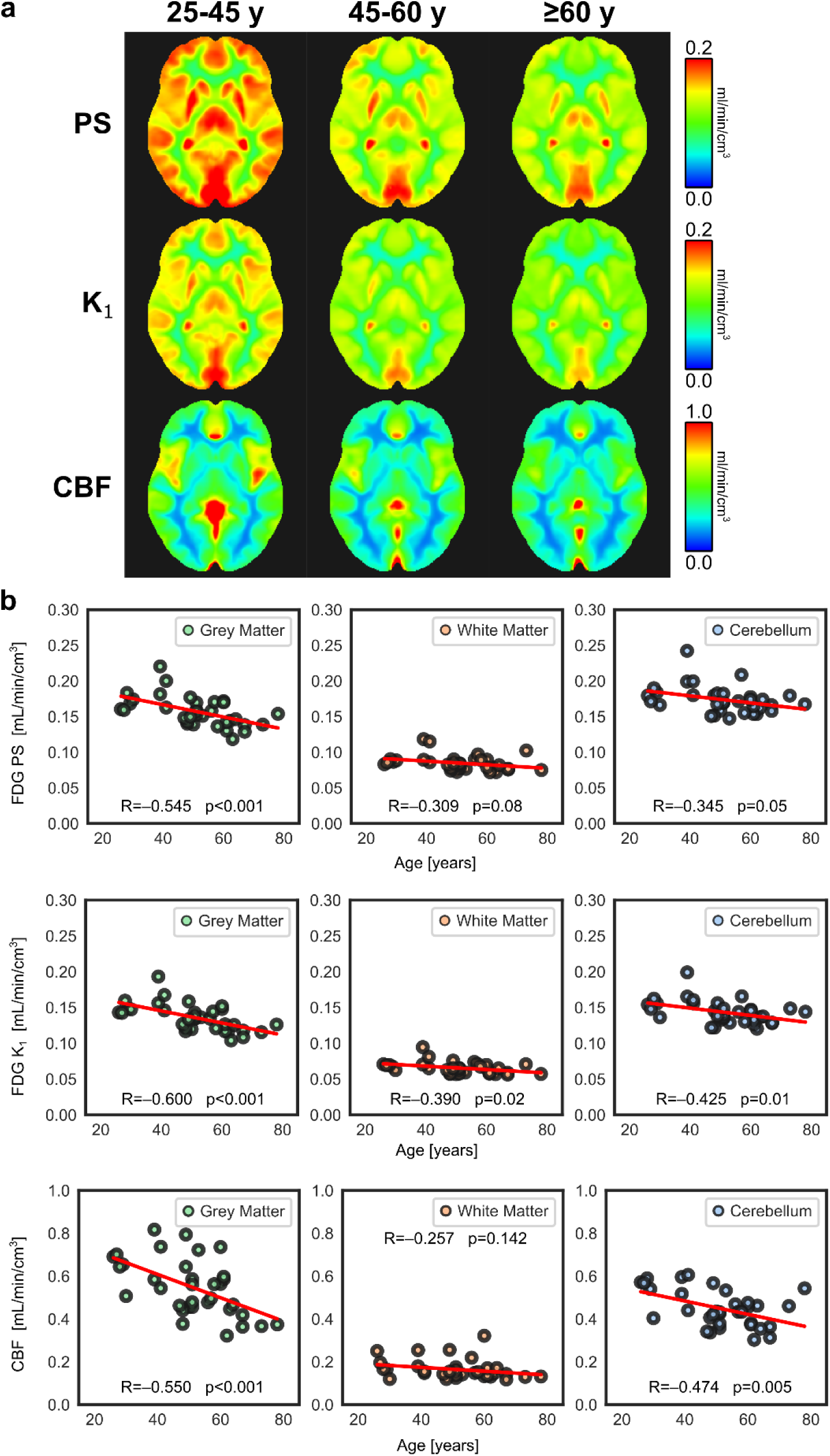
Blood-brain barrier (BBB) permeability-surface area (PS) product, BBB transport rate K_1_, and cerebral blood flow (CBF) in healthy aging with ^18^F-fluorodeoxyglucose (FDG) PET. **a**, Parametric images of FDG BBB PS, K_1_, and CBF non-rigidly registered to the MNI-152 space and averaged across healthy subjects in three age groups (25-45 y, N=9; 45-60 y, N=14; ≥60 y, N=11). **b**, Regional analysis shows significant decreases in FDG BBB PS in grey matter with age, while decreasing trends approached significance in white matter and cerebellum. FDG BBB transport K_1_ significantly decreased in all brain regions likely due to joint decrease of CBF and PS with age. PS, K_1_, and CBF are in units of ml/min/cm^3^.

To visualize inter-subject parametric averages across demographics, we non-rigidly transformed each subject’s brain parametric images into the Montreal Neurological Institute (MNI)-152 space^38,39^ and averaged across three age ranges (25-45 years, N=9; 45-60 years, N=14; ≥60 years, N=11). Inter-subject average parametric images showed progressively decreasing FDG BBB PS and K_1_ with age, particularly in the grey matter. Similar decreases in CBF were observed with age (Figure 4) as expected from prior work,^40^ but no significant associations were detected for FDG extraction fraction. Our non-invasive single-tracer method resolved multiparametric associations with age and may have utility in studies of the aging human BBB.

### Evaluating Brain-Body Crosstalk in Metabolic Dysfunction-Associated Steatohepatitis

We leveraged total-body dynamic PET and our ^18^F-FDG BBB permeability imaging method to explore brain-body crosstalk in systemic disease states. We studied metabolic dysfunction-associated steatotic liver disease (MASLD), the most common chronic liver disease globally^41^ with potential associations with cognitive impairment.^42^ However, there is a paucity of data on the involvement of the BBB in MASLD-related cognitive impairment in humans, especially at the molecular level. Here, we conducted the first human BBB study in MASLD with total-body dynamic FDG-PET, applying our FDG BBB PS method in thirty patients with biopsy-graded MASLD-related liver inflammation (i.e., MASH)^43^ and compared against thirteen age-matched healthy controls.

Parametric imaging of FDG BBB PS and regional analysis showed decreased FDG BBB PS in patients with severe hepatic lobular inflammation (N=17) compared to those with mild inflammation (N=13) and age-matched controls (N=13; Figure 5). Mean grey matter FDG PS was 0.145±0.025 ml/min/cm^3^ in the severe inflammation cohort, which was significantly lower than that of mild inflammation (0.165±0.017 ml/min/cm^3^; P=0.047) and age-matched controls (0.169±0.022 ml/min/cm^3^; P=0.013). Significant differences mostly persisted in white matter and cerebellum. Similarly, FDG BBB K_1_ significantly differed (P<0.01) between healthy controls and severe liver lobular inflammation groups in all brain regions of interest, but not between mild and severe inflammation groups except in the cerebellum (P=0.031). CBF did not significantly differ (P>0.05) between the three groups in any brain regions of interest, suggesting an effect on the BBB but not CBF in our cohort of MASLD patients with severe liver inflammation. Severe liver inflammation may therefore be a contributing factor to MASLD-related BBB dysregulation, possibly through proinflammatory cytokines circulating in blood and disrupting BBB transport.^4^

**Figure 5.**
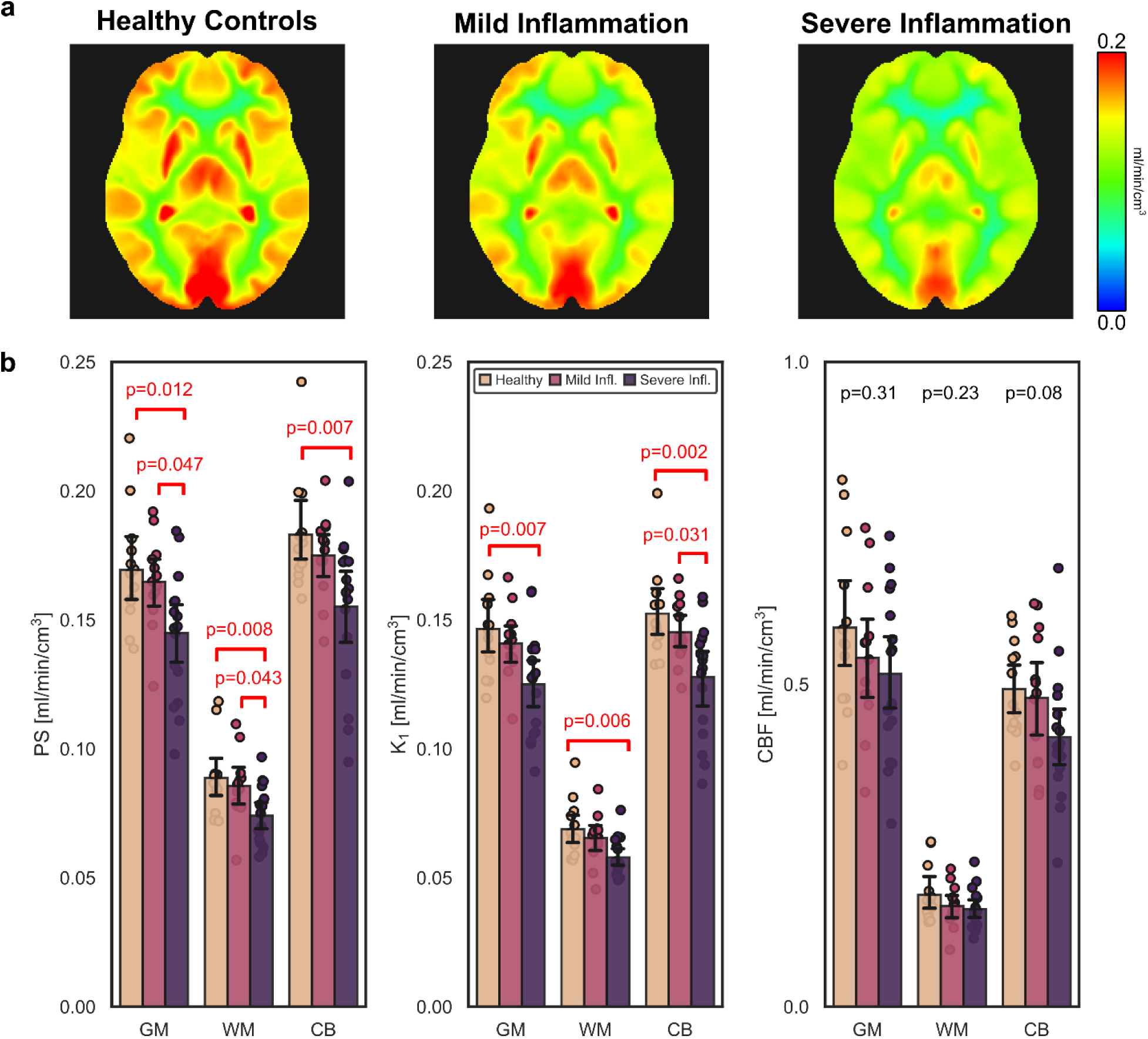
Blood-brain barrier (BBB) permeability-surface area (PS) product of ^18^F-fluorodeoxyglucose (FDG) in metabolic dysfunction-associated steatotic liver disease (MASLD). **a**, FDG BBB PS parametric images non-rigidly registered to the MNI-152 space and averaged across subjects grouped as age-matched controls (N=13) and patients with mild (N=13) and severe (N=17) MASLD-related lobular liver inflammation. The average FDG BBB PS of patients with severe lobular inflammation was significantly lower than that of mild inflammation and controls. **b**, Regional analysis also supported significant decreases in FDG K_1_ mainly between controls and severe inflammation, but no significant differences were observed with cerebral blood flow (CBF).

### BBB PS of ^18^F-FDG is Associated with Fasting Blood Glucose

Chronic hyperglycemia is known to downregulate glucose transporter 1 (GLUT1) expression at the BBB, leading to reduced BBB glucose transport.^44^ This may have significant clinical implications in diseases including diabetes mellitus and MASLD in which hyperglycemia is common. In our MASLD analyses, we also found that fasting blood glucose level was a significant covariate between the inflammation groups (P<0.001). Group-level comparisons of FDG PS were not significant after adjusting for blood glucose (P=0.279). Averaged inter-subject FDG BBB PS parametric images for three blood glucose ranges (normal, medium, and high) showed progressively lower FDG BBB PS with higher blood glucose levels (Figure 6a). Our data suggests that BBB dysregulation may be multifactorial or glucose mediated.

**Figure 6.**
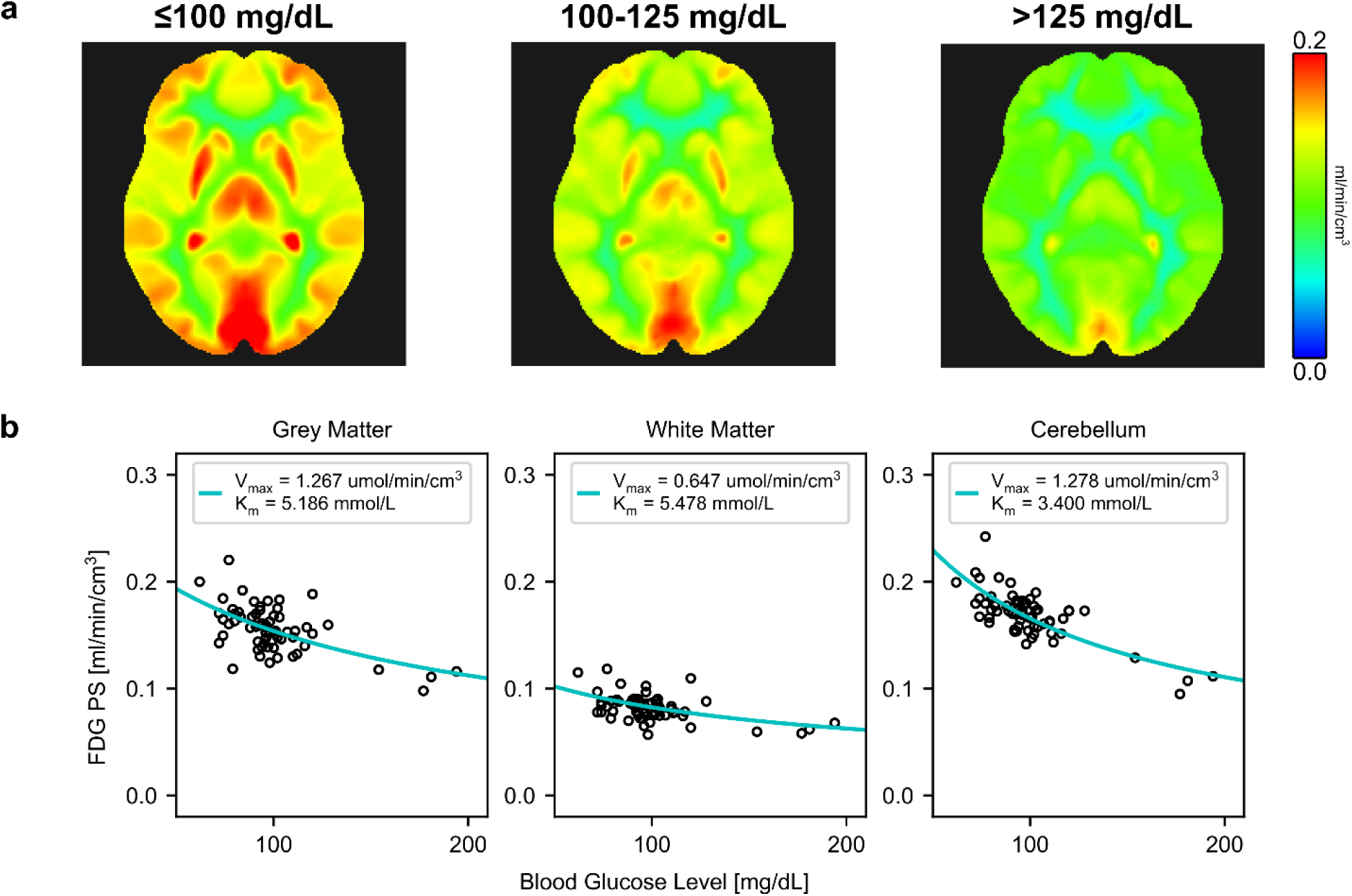
Blood-brain barrier (BBB) permeability-surface area (PS) product of ^18^F-fluorodeoxyglucose (FDG) and blood glucose level. **a**, FDG BBB PS parametric images averaged across blood glucose ranges showed that FDG PS decreased with higher blood glucose levels, with notable decreases at hyperglycemia (>125 mg/dL; N=5). **b**, Cohort-level Michaelis-Menten transporter kinetics across 64 analyzed subjects with total-body dynamic FDG-PET. PS is in units of ml/min/cm^3^.

To quantify the relationship between FDG BBB PS and blood glucose, we computed population-based Michaelis-Menten transporter kinetics^45^ by utilizing the range of blood glucose levels (62 to 194 mg/dl) across all 64 analyzed subjects with total-body dynamic FDG-PET (Figure 6b). The fitted maximal transport rate (V_max_) for FDG was 1.27, 0.65, and 1.28 µmol/min/cm^3^, in grey matter, white matter, and cerebellum, respectively, closely agreeing with preclinical estimates of FDG Michaelis-Menten kinetics^45^ and a human MR spectroscopy study showing brain glucose V_max_ is 2-3× greater in grey matter than that of white matter.^46^ The half-saturation constant (K_m_) ranged from 3.40 to 5.48 mmol/L between regions in general agreement with prior work.^45^ These data cross-validated the proposed HTR method for FDG BBB PS imaging. While it is difficult to measure regional blood glucose levels in the brain, FDG BBB PS provides detailed spatial information beyond a single blood glucose measure, possibly related to BBB transporter expression and function.

## Discussion

The BBB is the primary site of molecular exchange between the systemic circulation and brain parenchyma; however, *in vivo* molecular probing of the human BBB has thus far been limited by the lack of efficient translational imaging methods to specifically measure molecular BBB permeability. Herein, we developed a non-invasive multiparametric PET method to measure the molecular BBB PS of radiotracers with a single-tracer dynamic PET scan. We first demonstrated voxel-wise measurement of the molecular BBB PS of three PET tracers (^18^F-fluciclovine, ^18^F-FDG, ^11^C-butanol) spanning a wide range of BBB permeabilities. Focusing on FDG, we then demonstrated three clinical applications: BBB PS associations with age in healthy subjects, BBB dysregulation in MASLD-related liver inflammation, and an investigation of FDG BBB PS associations with blood glucose levels. Each of these studies has important implications in studying and characterizing healthy aging, brain-body crosstalk in chronic liver disease,^41^ and diabetes, respectively. The results collectively point to the critical need for *in vivo* molecular probing of the human BBB to elucidate the functional health of this dynamic barrier. We present a new paradigm to non-invasively study BBB function at the molecular level with a single dynamic PET scan.

Our non-invasive single-tracer method is a significant advancement over existing methods for imaging BBB PS. Past efforts with DCE-MRI have been limited to inert contrast agents with a low extraction fraction,^9,10^ mainly assessing the BBB as a structural barrier. The complexity of serial dual-tracer PET imaging^13–15^ has limited its widespread use in both preclinical and human imaging research^7,47^ despite the PS product prevailing as the most specific measure of BBB permeability.^6^ We show our method can measure BBB PS across three orders of permeability magnitude, opening opportunities to apply this method to the numerous molecular PET tracers already available for research and clinical use and which cross the BBB through a diverse set of transport mechanisms.

The proposed method was enabled by HTR dynamic imaging in combination with advanced kinetic modeling for joint estimation of CBF and tracer-specific BBB transport rate. Such a HTR method was challenging, if not impossible, using past-generation PET scanners due to their poor temporal resolution, insufficient statistical quality of dynamic data, and lack of a reliable image-derived input function.^23^ Though our first demonstration of molecular BBB PS imaging was performed with an advanced total-body scanner, the clinical and research adoption of this and similar high-sensitivity scanners^21,48–50^ is rapidly growing with over sixty installations worldwide. Advances in brain-dedicated PET imagers^50^ and image reconstruction methods^51,52^ are also imminent, bringing higher spatial and temporal resolution for dynamic imaging to enable our HTR kinetic modeling method into broader settings.

The interpretation of the BBB PS depends on the specialized molecular transport mechanism of the tracer and the vascular environment.^1^ For example, ^11^C-butanol freely diffuses across the BBB, leading to an extraction fraction of ≈100% as previously suggested^16,23^ and confirmed with our method. For ^18^F-fluciclovine and ^18^F-FDG, BBB transport is passively facilitated primarily by the sodium-independent L-type large neutral amino acid transporter 1 (LAT1)^53^ and glucose transporter 1 (GLUT1),^54^ respectively. The greater BBB PS of ^18^F-FDG over ^18^F-fluciclovine can partially be explained by the two-order of magnitude greater expression of GLUT1 found over LAT1 in a post-mortem proteomic study in humans.^55^ Further quantitation with Michaelis-Menten transporter kinetics could explain differences in molecular PS for facilitative transport.^45^ In comparison, DCE-MRI measures of BBB permeability mainly represent non-specific leakage of contrast material associated with BBB breakdown^8–10^ as gadolinium contrast agents are not known to cross an intact BBB effectively.^5^ Differences in BBB transport mechanisms may explain why BBB permeability changes in different ways for each tracer. DCE-MRI-derived PS was shown to increase with age due to increased vascular leakage^8^ while our healthy aging study showed that FDG BBB PS decreases with age possibly due to reduced GLUT1 transporter expression.^56^

Multiparametric imaging of CBF, molecular BBB permeability, and transport can augment a radiotracer’s standard use, opening opportunities otherwise challenged by the complexity of multi-tracer imaging. For instance, impaired CBF, dysregulated BBB permeability and transport, and reprogrammed cellular metabolism (using the standardized uptake value or net uptake rate, K_i_),^18^ which are common markers of neurovascular dysfunction,^2^ can now be efficiently assessed with our multiparametric imaging method from a single dynamic FDG-PET scan. The prevalence of FDG-PET in oncology may accordingly be opportune to study cancer-related cognitive impairment^57^ with our multiparametric brain imaging method. Beyond ^18^F-FDG, our single-tracer method efficiently can add multiparametric depth for studying Alzheimer’s disease with amyloid^58^ and tau^59^ radiotracers (e.g., ^18^F-florbetaben and ^18^F-PI-2620, respectively), synaptic density in major depression with radioligands (^11^C-UCB-J),^60^ and the neuroimmune system with ^18^F-DPA-714 for neuroinflammation^61^ or ^18^F-AraG for imaging T-cell activation.^62^

A major limitation of this work is the lack of ground truth values in humans for validation of our PS measurements. This in part reflects the practical difficulties of measuring the molecular BBB PS of radiotracers in humans using existing methods. However, several of our results characterized the proposed method indirectly. First, the measured CBF was consistent across three very different radiotracers and were all comparable to population-based values reported in the literature.^16,17,23^ The method was also able to accurately estimate an extraction fraction equal or close to 100% for the freely diffused tracer ^11^C-butanol^16^ and a small extraction fraction for ^18^F-fluciclovine, the latter known to have low uptake in the brain.^34,63^ Second, FDG BBB PS in healthy subjects was also comparable to those reported in the literature by other methods.^12,13^ Our observed negative association of FDG BBB PS with age is concordant with evidence that the expression of GLUT1 at the BBB decreases with age.^56^ Third, we also used the FDG BBB PS estimates to derive its theoretical Michaelis-Menten transporter kinetics, which agreed with those derived from preclinical and human data.^45,46,64^ These results increase confidence in our proposed method.

This study also had other limitations. It is possible that our PS estimates not only comprise molecular transport through the BBB, but also through parenchymal cell membranes. This challenge persists in other methods^7,12,65^ and we mitigated this by studying only the first two minutes of the dynamic scan. Our PS estimates may therefore be marginally overestimated. Future work will comprise further optimization of scan duration and extension of the HTR kinetic model from one tissue compartment to two tissue compartments to better capture the full kinetics of metabolic tracers like ^18^F-FDG. In addition, our studies of age, MASLD, and blood glucose were exploratory and not specifically designed to answer a biological hypothesis. For example, our pilot study of MASLD showed an association of FDG BBB PS with liver inflammation but the result may be confounded by blood glucose. Liver inflammation, insulin resistance, and diabetes may collectively contribute to the dysregulation of the BBB^3,4,66^ and the complex multivariate interactions could not be fully resolved with our relatively small sample size. Our main aim was instead to showcase the potential of our novel method. Future studies with additional controls and complementary data will better elucidate the biological and clinical significance of the molecular BBB PS. Finally, the proposed method involves radiation exposure. However, the maximum exposure (7.0 mSv) in our studies is well within the acceptable range for healthy subjects, as compared to the average annual natural background radiation of 3.1 mSv in the United States. Furthermore, future algorithm improvements (e.g., deep learning-based image reconstruction^52^) may help reduce radiation dose in the future.

The proposed method has many potential applications beyond the demonstrations in this paper. In drug development, PS remains the key parameter describing drug permeability across the BBB^6,7^ and our method may revitalize its adoption in both preclinical drug development studies and in human studies. Broader quantification of molecular permeability may also support the development of *in silico* methods for drug delivery and discovery.^7,47,65^ Beyond the brain, quantifying vascular permeability may add a new dimension to study cardiovascular disease,^67^ design treatment delivery systems, and monitor the vascular toxicity of systemic cancer therapies.^68^ Furthermore, changes in gut vascular permeability have been observed due to pathogenic bacteria^69^ and gut-brain interactions have shown a dysregulation of the BBB in mice lacking gut microbiota.^70^ Measuring molecular permeability at systemic capillaries such as at gut vasculature,^69^ liver sinusoids,^71^ and the blood-tumor barrier^72^ may require further development as transport mechanisms likely differ relative to the highly controlled BBB.^1^ Such methodological advances in combination with total-body PET may enable vascular permeability studies along the brain-body axis, with potential applications to design and monitor the delivery of systemic therapies. Thus, our developed method may serve as a powerful translational framework to study the role of molecular barrier function in neurological and systemic diseases.

## Methods

### Study Design

The primary objective of this study was to develop a single-tracer method of quantifying and imaging the BBB PS of PET radiotracers and to demonstrate its importance in characterizing molecular BBB permeability. To this end, this study was divided into five experiments. The first experiment focused on demonstrating the need and capability of high temporal resolution (HTR) dynamic imaging and more advanced kinetic modeling enabled by total-body PET to jointly estimate CBF and tracer-specific BBB transport rate K_1_. The second experiment was to use CBF and K_1_ to quantify and image differences in BBB PS between PET radiotracers. We included HTR dynamic PET studies scanned with three radiotracers thought to encompass a wide range of BBB PS due to their previously reported BBB transport rate values.^12,16,34^ In the third experiment, we investigated the molecular BBB PS of ^18^F-FDG and its association with aging. In the fourth experiment, we conducted an exploratory analysis of FDG BBB PS to investigate a potential brain-body crosstalk in patients with metabolic dysfunction-associated steatotic liver disease (MASLD) enrolled for an imaging trial of liver inflammation. Lastly, the relationship between FDG BBB PS and fasting blood glucose was investigated using the pooled healthy subjects and MASLD patients.

### High-temporal Resolution Dynamic Imaging with Total-body PET

Total-body positron emission tomography (PET) enabled non-invasive measurement of the arterial input function from a major blood pool (e.g., the ascending aorta) and HTR dynamic imaging for single-tracer parametric imaging of BBB PS. All participants were scanned on the uEXPLORER total-body PET/CT system (United Imaging Healthcare) with a 194-cm axial field of view and exceptional detection sensitivity for high spatial resolution (≈3.0 mm full-width at half maximum resolution by the NEMA standard)^21^ and HTR dynamic imaging. The performance characteristics and potential of the uEXPLORER for HTR dynamic imaging have been reported in prior studies.^21,73,74^ All participants received either an ultra low-dose or low-dose total-body CT (140 kVp with dose modulation at 5 or 50 mAs maximum tube current-exposure time product, respectively, corresponding to effective doses of ≈1 mSv or ≈10 mSv) for attenuation correction and anatomical localization. Dynamic PET imaging commenced immediately prior to bolus injection of the radiotracer. For brain kinetic modeling of each dynamic PET scan, we performed HTR reconstructions of the first two minutes (framing: 60×1 s, 30×2 s) using vendor-provided reconstruction software and standard corrections for attenuation, scatter, randoms, dead time, and decay.^21^ Specifically, a time-of-flight ordered subset expectation-maximum algorithm with 4 iterations and 20 subsets was used to reconstruct each dynamic image. We pooled total-body dynamic PET scans from several human studies with institutional review board approval and written informed consent. The effective dose of PET scans varied depending on the radiotracer and injected activity. For the investigated radiotracers in this study, approximate effective doses per unit activity were 19 µSv/MBq for ^18^F-FDG, 22 uSv/MBq for ^18^F-fluciclovine, and 4 µSv/MBq for ^11^C-butanol,^11^ resulting in a mean effective dose of 7.0 mSv, 6.8 mSv and 1.1 mSv for the three studies, respectively.

### High-Temporal Resolution Kinetic Modeling and Measuring BBB PS

The early kinetics of a radiotracer in the brain were quantified by using the adiabatic approximation to the tissue homogeneity (AATH) model^30^ applied on the first two minutes of HTR dynamic PET data. The AATH model offers a closed-from time-domain solution to a distributed kinetic model comprised of a spatiotemporally distributed intravascular space and a compartmental extravascular space.^30^ The impulse response function of the AATH model is (Supplementary Figure 2a)

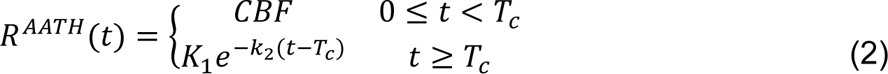

Where *CBF* is the cerebral blood flow (ml/min/cm^3^), *T*_*c*_ is the mean vascular transit time (s), *K*_1_ is the BBB transport rate (ml/min/cm^3^) of the radiotracer, *k*_2_ is in BBB clearance rate (min^-1^). This solution describes a vascular phase (0 ≤ *t* < *T*_*c*_) during which time the tracer traverses the intravascular space while permeating to the extravascular space. The tissue phase (*t* ≥ *T*_*c*_) follows and describes the return of extracted tracer to the intravascular space and subsequent venous clearance.

For a general arterial input, *C*_*a*_(*t*), the tissue time-activity curve is

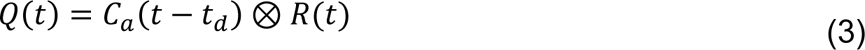

where *t*_*d*_ is a time delay parameter accounting for the time difference between tracer arrival at the ascending aorta and the regional cerebral artery. A parametric form of the AATH time-activity curve can be derived by substituting (2) into (3):

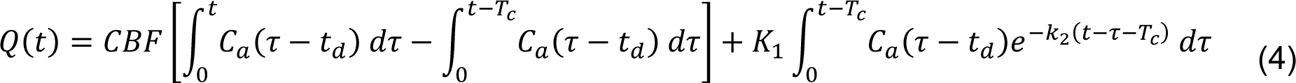

We used a basis function method to estimate the five parameters of the time delay-corrected AATH model. Specifically, we performed a naïve grid search of *t*_*d*_ ∈ [0,16] s and *T*_*c*_ ∈ [3, 16] s at 0.25 s intervals and used 100 logarithmically spaced *k*_2_ ∈ [0.006, 3] min^-1^. A non-negative linear least squares algorithm was then used to estimate *C* and *K*_1_.^75^ Based on time-delay and mean vascular transit time estimates from regional kinetic analysis, we reduced the grid search interval of *t*_*d*_ and *T*_*c*_ to 0.5 s to reduce computation time for voxel-wise parametric imaging. We interpreted the CBF term as the intravascular basis and the *K*_1_ term as the extravascular tissue basis to resolve the distributions of the fitted time-activity curve.

The standard one-tissue compartment (S1TC) model was used for comparison against existing methods. The impulse response function of the S1TC model is (Supplementary Figure 2b)

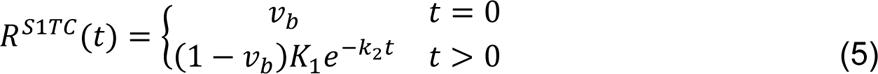

where *v*_*b*_ is the fractional blood volume (ml/cm^3^). The S1TC differs from the AATH model as it assumes instantaneous distribution of tracer in the intravascular space and neglects the finite transit time required for tracer to traverse the blood vessel volume. As such, the S1TC response function lacks a finite-length vascular phase and the model describes that tracer is immediately cleared from tissue at *t* > 0. A parametric form of the S1TC time-activity curve can be derived by substituting (5) into (3)

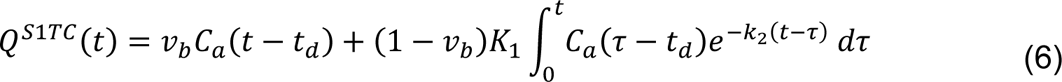

The four parameters of the time delay-corrected S1TC model were estimated in a manner similar to the AATH model by a basis function method where *v*_*b*_ and (1 − *v*_*b*_)*K*_1_ were estimated by a non-negative linear least squares algorithm.^75^ For all investigated tracers and for both AATH and S1TC methods, we assumed an absence of metabolites^17,34,63^ and that the whole-blood tracer activity was equal to that in blood plasma over the first two-minutes of the dynamic PET scan.

An advantage of the AATH model^30^ and other distributed models^24^ is their ability to jointly estimate CBF and the tracer-specific BBB transport rate K_1_ from HTR dynamic PET data whereas the S1TC model can only estimate K_1_. The PS product (Eq.(1)) can then be calculated from CBF and K_1_ by rearranging the Renkin-Crone equation^32,33^

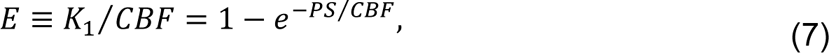

where *E* is the extraction fraction.

### Imaging the Molecular BBB PS of Different PET Tracers

To demonstrate that our method can measure across different radiotracers with a wide range of BBB permeabilities, we included fifteen age-matched participants scanned with one of either ^18^F-fluciclovine, ^18^F-FDG, or ^11^C-butanol (N=5 each). These three radiotracers were chosen to span a wide range of low to very high BBB PS as expected from their previously reported BBB transport rate values.^12,16,34^ ^18^F-fluciclovine is a radiolabeled analogue of leucine, an essential amino acid, which has demonstrated low brain uptake and BBB transport rate on the order of 10^-2^ ml/min/cm^3^.^34,63^ ^18^F-FDG is a glucose analogue with moderate BBB PS on the order of 10^-1^ ml/min/cm^3^.^12,13^ ^11^C-butanol is a lipophilic alcohol and considered a favorable flow radiotracer due to its predictably high extraction fraction of ≈100% owing to its free apparent diffusion across the BBB.^16,23^ ^18^F-fluciclovine, ^18^F-FDG, and ^11^C-butanol PET studies were scanned using a mean (±standard deviation) activity of 309±8 MBq (range: 298 to 318 MBq), 370±16 MBq (349 to 395 MBq), and 282±10 MBq (267 to 296 MBq). The mean age of the participants was 63±6 years (range: 54 to 73 years) and there were no significant differences in age between tracer groups (P=0.796). Thirteen of fifteen participants were male, and both female participants were scanned with ^11^C-butanol. Most participants were healthy volunteers except those scanned with ^18^F-fluciclovine, who had biochemically recurrent prostate cancer.

### Model Comparison, Practical Identifiability Analysis, and Statistical Analysis

To assess the need for HTR dynamic imaging, our original measured regional time-activity curves were frame averaged at 1 to 10 s intervals and fitted with the AATH and S1TC models. The Akaike Information Criterion (AIC)^31^ was computed to statistically determine which model produced a better fit at different temporal resolutions. A lower AIC indicated better statistical fit after adjusting for the trade-off between model complexity and residual model fitting error. The AIC was computed as

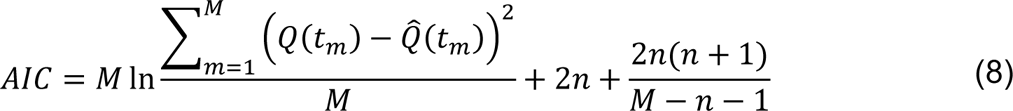

where *Q*(*t*) is the measured time-activity curve, *Q̂*(*t*) is the model fitted time-activity curve, *M* is the number of time frames, *t*_*m*_ is the midpoint time of the *m*th frame, and *n* is the number of model parameters (*n* = 5 for AATH, *n* = 4 for S1TC).

Practical identifiability analysis was conducted to assess the reliability of our parameter estimates.^76^ For each subject’s AATH fitted curves, 1024 realizations of time-varying noise were added and AATH parameters were estimated from these simulated noisy time-activity curves. Parametric error mean and standard deviation were computed across the 1024 noise realizations. We reported error mean and standard deviation for each radiotracer group and brain region. Time-varying noise accounted for frame duration, radionuclide decay, and time-varying activity concentration, and the noise was scaled by the estimated standard deviation of the normally distributed fitting residuals as described in prior work.^76^ Additional simulations were conducted to determine the identifiability of PS when varying extraction fraction. Using an average ascending aorta curve determined from our dataset, we simulated tissue time-activity curves using the AATH model with fixed CBF = 0.50 ml/min/cm^3^, t_d_ = 2 s, T_c_ = 4.5 s as guided by our whole-brain data. Extraction fraction E was varied from 1% to 99% and extravascular distribution volume V_e_ was also varied from 0.25 to 1.0 ml/cm^3^ from which k_2_ = K_1_ / V_e_ was derived. A noise scale factor of 4.8 were used as estimated from the measured regional time-activity curves as described.

Subsequent statistical analyses were performed with IBM SPSS Statistics 29 using a two-tailed alpha of 0.05 for statistical significance. A one-way analysis of variance (ANOVA) with post hoc Bonferroni-corrected pairwise comparisons was used to compare differences in means between groups with more than two categories. A t-test was used to compare the molecular BBB PS of FACBC and FDG due to the absence of PS values for butanol.

### FDG BBB PS in Aging

To investigate the association of FDG BBB PS with age, we included thirty-four healthy subjects (21 females, 13 males; mean age: 51±13 years, range: 26 to 78 years) who received 60-minute total-body dynamic FDG-PET (mean activity: 358±33 MBq). We explored associations between age and FDG BBB PS, K_1_, and CBF by the Pearson coefficient and linear regression. Linear regression slopes were reported to indicate parametric change per year change in age. Percent changes were computed by regressing age with the logarithm of the parameter. We compared parametric images across three age groups (25-45 years, N=9; 45-60 years, N=14; 60 years, N=11), aiming for a similar number of subjects in each group.

The five FDG-PET participants from the three-radiotracer experiment were a subset of those from this experiment. Subjects had no history of major disease within the last five years or ongoing acute inflammation. Participants fasted for at least 6 hours prior to FDG-PET (mean: 11±2 hours) and had a mean fasting blood glucose level of 91±12 mg/dl (range: 62 to 116 mg/dl). The mean body-mass index (BMI) was 28.0±5.6 kg/m^2^ (range: 17.5 to 37.6 kg/m^2^) with approximately equal distributions in the number of subjects with healthy weight (18.5 to 24.9 kg/m^2^; N=11), overweight (25.0 to 29.9 kg/m^2^; N=10), and obesity (≥30 kg/m^2^; N=12).

### FDG BBB PS in MASLD

This exploratory analysis of BBB permeability in systemic disease included 30 consecutive patients receiving liver biopsy and 60-minute total-body dynamic FDG-PET (mean activity: 186±13 MBq) between July 2020 and February 2023. Liver biopsy and FDG-PET were obtained within a median of 5.0 (interquartile range, IQR: 2.1 to 11.6) weeks of one another. An expert pathologist graded the liver biopsies by the MASLD activity score (MAS; ranging from 0 to 8 where a higher value indicates greater severity of MASLD), equal to the sum of sub-component scores for steatosis (0 to 3), lobular inflammation (0 to 3), and ballooning degeneration (0 to 2).^43^ Patients fasted for at least 6 hours prior to FDG-PET.

Here, we dichotomized patients into mild and severe lobular inflammation biopsy scores (e.g., mild included scores 0 and 1, and severe included scores 2 and 3) and their FDG early brain kinetic parameters were compared. To serve as a healthy control group, we also included 13 age-matched healthy subjects from the thirty-four participants described in the healthy aging experiment. These participants were assumed to have no abnormal liver findings as supported by qualitative readings of their FDG-PET and CT scans by a nuclear medicine physician. The mean age was 49.6±12.5 y, 52.4±13.0 y, and 51.0±11.0 y in the healthy control, mild lobular inflammation, and severe lobular inflammation groups, respectively, and did not significantly differ by a one-way ANOVA (P=0.84). We used the univariate general linear model tool in SPSS for multivariable regression analysis to adjust for blood glucose level when comparing FDG brain kinetics between age-matched healthy controls (mean blood glucose level: 90.0±10.8 mg/dl; N=13), mild lobular inflammation (98.0±15.8 mg/dl; N=13), and severe lobular inflammation (118.6±35.8 mg/dl; N=17). We also examined groupings by fasting blood glucose levels between 70 to 100 mg/dl (normoglycemia; N=25), 100 to 125 mg/dl (N=13), and >125 mg/dl (hyperglycemia; N=5).

### Michaelis-Menten Transporter Kinetics with FDG BBB PS

To cross-validate our FDG BBB PS estimates against literature values, we computed Michaelis-Menten transporter kinetic parameters across all total-body dynamic FDG-PET scans analyzed in our study. A total of 64 subjects were included of whom 34 were healthy subjects and 30 were patients with MASLD as described previously. Blood glucose levels were measured by a fingerstick test prior to FDG-PET and ranged from 62 to 194 mg/dl in our cohort. We used a non-saturable Michaelis-Menten facilitative transporter kinetics model described previously:^45^

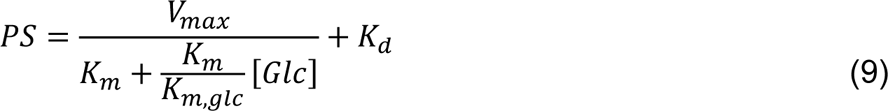

where *V_max_* is the maximal transport rate (µmol/min/cm^3^) of FDG, *K*_*m*_ is the half saturation constant (mmol/L) of FDG, *K*_*m*_/*K_m,glc_* is the ratio of half saturation constants between FDG and glucose, and [*Glc*] is the blood glucose concentration (mmol/L) in competitive transport with FDG, and *K*_*d*_ is the non-saturable transport rate. A Levenberg-Marquardt algorithm with a maximum of 100 iterations was used to solve for *V_max_*, *K*_*m*_, and *K*_*m*_/*K_m,glc_* using initial parameters 1.0 µmol/min/cm^3^, 5.0 mmol/L, and 1, respectively. Due to the relatively narrow range of blood glucose levels available in this study, we fixed *K*_*d*_ to 0.022 ml/min/cm^3^ based on prior data.^45^

### Image Analysis

An image-derived input function was delineated from the ascending aorta for kinetic analysis of all radiotracers. For regional kinetic analysis, additional regions of interest in the grey matter, white matter, and cerebellum were manually segmented to extract average regional time-activity curves. Regions of interest were delineated using 3D Slicer^77^ and by referring to a combination of dynamic and static PET frames and the attenuation correction CT. Cerebellar grey and white matter were not explicitly distinguished in our segmentations. Large cerebral vessels such as at the Circle of Willis and sagittal sinus were avoided.

Voxel-wise parametric imaging was performed with the basis function method on the reconstructed dynamic images of 2.344-mm isotropic voxels. Dynamic images and generated parametric images were smoothed using the post-reconstruction kernel method, which is equivalent to a type of nonlocal means noise reduction.^78,79^ The kernel matrix was built for each PET scan from four composite image priors derived from the full dynamic study and 49-nearest neighbours within a 9×9×9 voxel space. For visualization, parametric images were aligned to the Montreal Neurological Institute (MNI)-152 space^38,39^ using the nifty_reg registration toolbox.^80,81^ For registration, two-minute static PET images were cropped to the brain and were used to compute rigid and affine transformations to the MNI-152 space. These computed transformations were then used to resample the parametric images to the MNI-152 space. Deformable registration was only used when generating group-averaged parametric images. For ^11^C-butanol, which often had K_1_ estimates equal to CBF, PS maps were generated by clipping extraction fraction values to 99.9% to avoid indeterminate outputs. This was only for visualization purposes and we accordingly did not quantify the PS of ^11^C-butanol.

## Data Availability

All data produced in the present study are available upon reasonable request to the authors.

## Acknowledgments

The authors gratefully acknowledge Dr. Elizabeth Li for her assistance in collecting the ^11^C-butanol PET data, Dr. Karen Matsukuma for her assistance in collecting histopathological data for the MASLD patient cohort, and the technologists and staff at the EXPLORER Molecular Imaging Center, particularly Lynda E. Painting, for their assistance in patient consent and data acquisition. This research was supported in part by National Institutes of Health grants R01 EB033435, R01 DK124803, and R01 CA206187. Image data in this work were also acquired under the support of In Vivo Translational Imaging Shared Resources with funds from National Institutes of Health P30 CA093373.

## Author contributions

G.W., K.J.C., and S.R.C. conceived the concept and designed the study. G.W. and Q.T. conducted the initial test. K.J.C. developed the data analysis pipeline, conducted the study, and analyzed the results. G.W. and S.R.C. jointly supervised the work. K.J.C. wrote, and G.W. and S.R.C. revised the first manuscript draft. K.J.C., Y.G.A., and B.A.S. reconstructed the PET data. T.J., R.D.B., and Y.G.A. contributed to data interpretation. Y.G.A., B.A.S., L.N., R.D.B, M.S.C., S.S., V.M., and V.L. contributed to the acquisition of the data. G.W., S.R.C., and R.D.B. obtained funding. G.W., S.R.C., and K.J.C. are the guarantors of the integrity of the entire study. All authors edited and approved the submitted version of the manuscript.

## Supplementary Materials

**Supplementary Table 1.** Practical identifiability analysis of molecular BBB transport kinetics of three radiotracers.

| Parameter | Mean ± Standard Deviation [%] | | |
| --- | --- | --- | --- |
|  | <sup>18</sup>F-fluciclovine | <sup>18</sup>F-FDG | <sup>11</sup>C-butanol |
| PS | 0.2±11.5 | −0.1±3.4 | — |
| CBF | 1.8±12.2 | 1.2±11.5 | 2.0±8.3 |
| K<sub>1</sub> | 0.2±11.3 | −0.1±2.2 | −0.1±2.0 |
| E | −0.5±13.9 | −0.1±10.8 | −1.5±6.3 |

Error percent mean and standard deviation of the permeability-surface area (PS) product, blood-brain barrier transport rate K<sub>1</sub>, cerebral blood flow (CBF), and radiotracer extraction fraction (E) for the three investigated radiotracers, <sup>18</sup>F-fluciclovine, <sup>18</sup>F-fluorodeoxyglucose (FDG), and <sup>11</sup>C-butanol. The identifiability of PS could not be computed for <sup>11</sup>C-butanol due to its median extraction fraction of 100%, which leads to indeterminate PS values (Eq. (1)). A positive value indicates our method overestimated the true value.

**Supplementary Figure 1.**
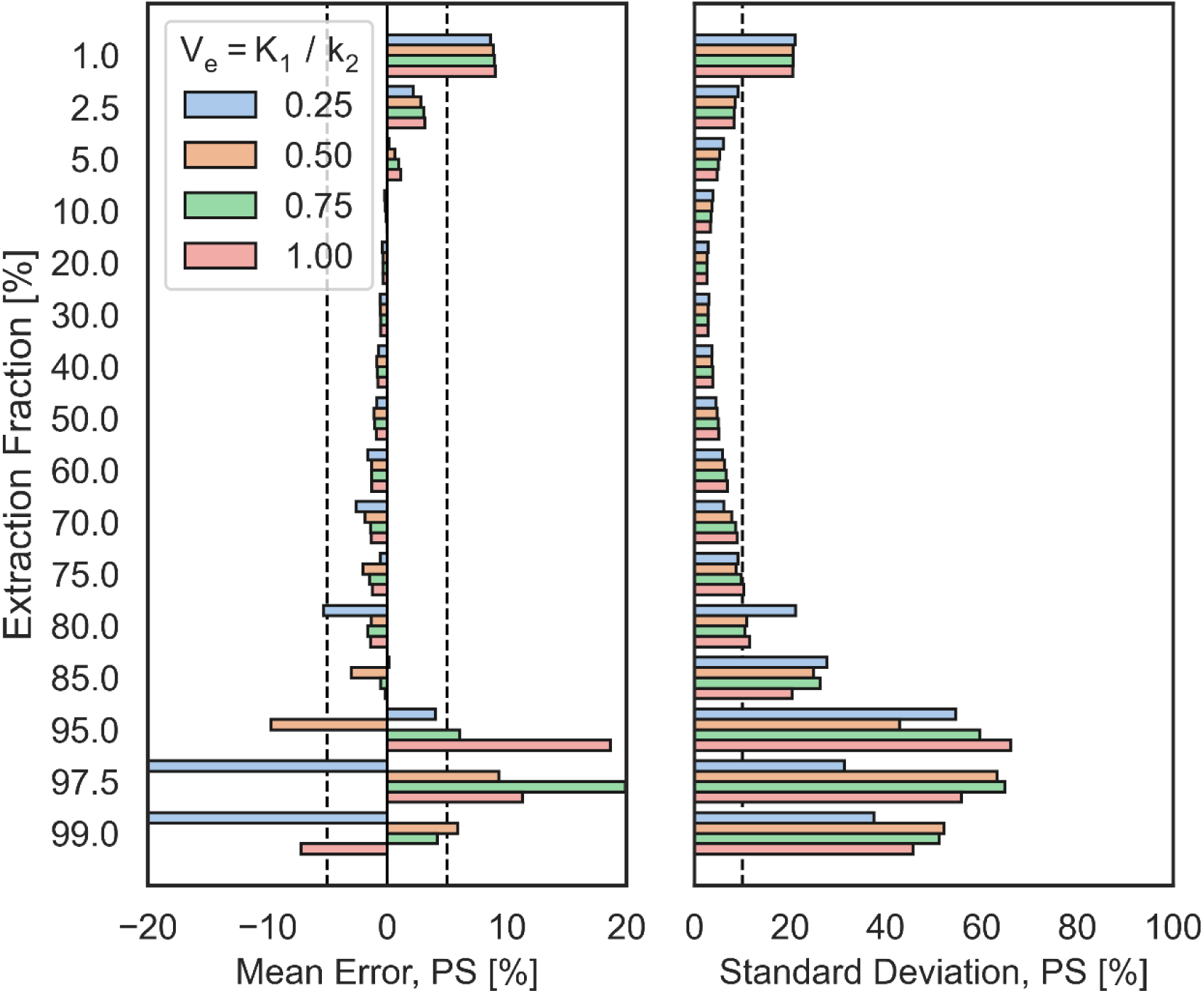
Practical identifiability analysis of the permeability-surface area (PS) product with respect to extraction fraction and extravascular distribution volume (V_e_).

**Supplementary Figure 2.**
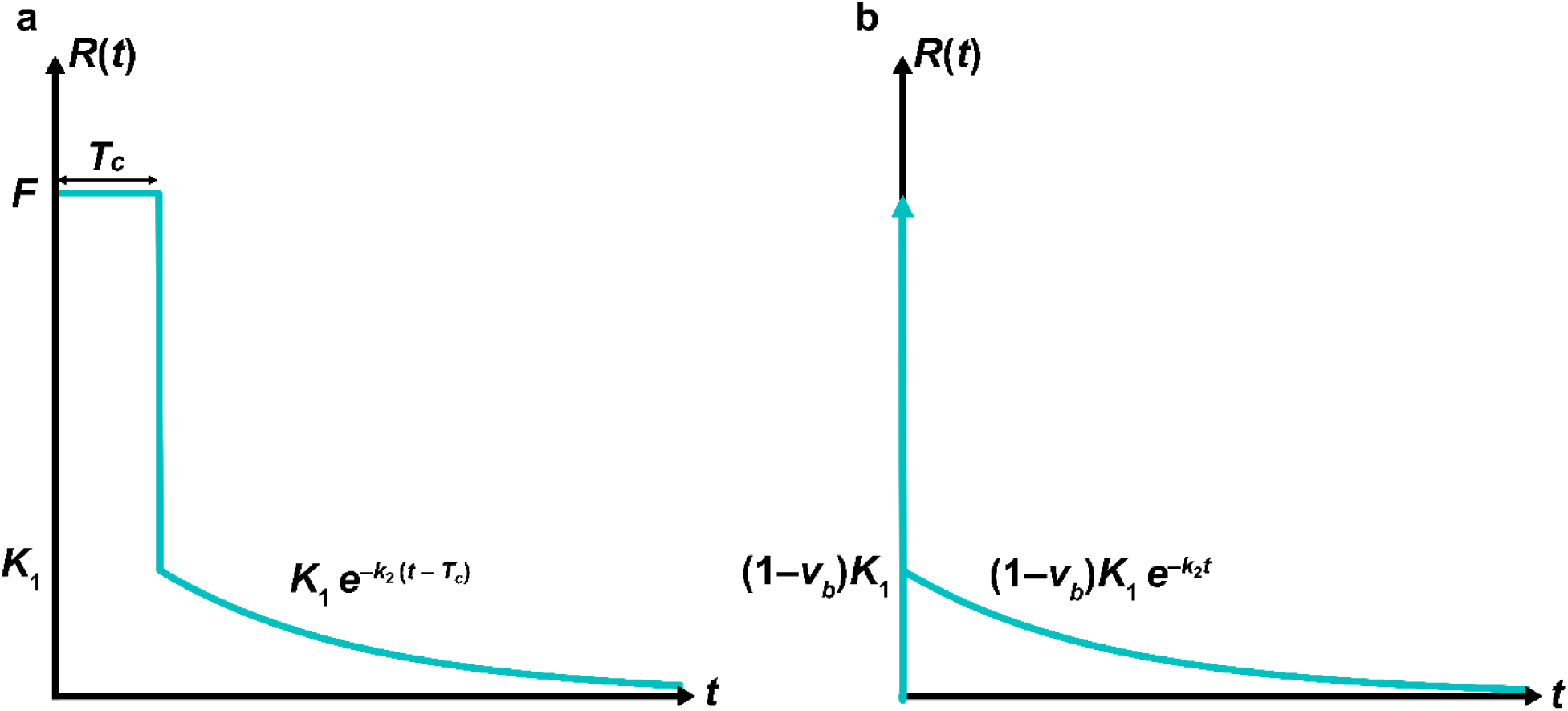
Impulse response functions (R(t)) of tracer kinetic models investigated in this study for modeling the early-dynamic PET data of a radiotracer. **a**, The adiabatic approximation to the tissue homogeneity (AATH) model. **b**, The standard one-tissue compartment model. The arrow represents a delta function with an area equal to the blood volume, v_b_. F indicates cerebral blood flow; K_1_, the blood-brain barrier (BBB) transport rate; k_2_, BBB clearance rate; T_c_, vascular mean transit time.

